# A Resistance Persistence Index (RPI) for antimicrobial resistance in wastewater

**DOI:** 10.1101/2024.11.06.24316840

**Authors:** Michael Hajkowski, Benjamin Rafael-Mingoa, Gabriela Crystal-Franco, Katherine Dick, Michael Acholonu, Sanjiev Nand, Luzmaria Soto, Brennan Nuque Withers, Morgan Hernandez, Ariel Cheng, Archana Anand

## Abstract

Antimicrobial resistance (AMR) represents a critical global health challenge, exacerbated by the proliferation of resistance genes in wastewater environments. This study introduces a novel Resistance Persistence Index (RPI) that integrates existing risk assessment frameworks with persistence and mobility metrics of antimicrobial resistance genes (ARGs) in wastewater treatment plant (WWTP) influent. We evaluated samples collected across wet and dry seasons, focusing on the dynamics of ARG diversity, abundance, and their associations with human impact. Our findings indicate a significant increase in unique ARGs during the wet season, with “antibiotic inactivation” as the predominant resistance mechanism. Using Zhang et al.’s ranking framework, we assessed the risk levels of various ARGs, revealing that while fewer high-risk ARGs were identified, they demonstrated substantial abundance. The RPI provided insights into how the combined effects of persistence and mobility influence the risk associated with different ARG families. This integrated framework offers a comprehensive tool for understanding and managing AMR in environmental settings, guiding future surveillance and intervention strategies.

## 1. Introduction and Background

Antimicrobial resistance (AMR) represents one of the most significant public health challenges of the 21st century, compromising the efficacy of antibiotics and leading to increased morbidity, mortality, and healthcare costs globally (World Health Organization, 2020; Centers for Disease Control and Prevention, 2021). Wastewater treatment plants (WWTPs) are increasingly recognized as critical reservoirs and dissemination points for antimicrobial resistance genes (ARGs) and antibiotic-resistant bacteria (ARB; Cacace et al., 2019; Rizzo et al., 2013; Michael et al., 2021). WWTP influent, which collects a variety of wastewater streams including domestic, hospital, and industrial effluents, acts as a complex environment where resistant microorganisms and antibiotics converge. Consequently, WWTP influent has become a valuable target for AMR surveillance and understanding the dynamics of resistance spread within environmental settings (Manaia et al., 2018; Riquelme et al 2022; Honda et al 2023).

A number of studies have explored AMR within WWTPs, with a focus on both the characterization and the quantification of ARGs and ARB (Liguori et al 2022; Ferraro et al 2024). Conventional methodologies include culture-based techniques to isolate specific resistant bacteria (Young et al 2016), quantitative polymerase chain reaction (qPCR) for ARG quantification (Schwartz et al 2003; {ruden et al 2006), and metagenomic sequencing to examine the resistome of influent samples. (Hendriksen et al 2019). These methods offer varying levels of insight, ranging from the identification of resistance phenotypes to an in-depth understanding of the diversity and abundance of ARGs across bacterial populations. However, each method has its own limitations. Culture-based techniques are often limited by their inability to capture the full diversity of resistant bacteria, as many environmental microorganisms are not easily culturable (Rizzo et al 2013). qPCR, while highly sensitive, is restricted to targeting known ARGs and cannot provide insight into novel resistance elements (Ishii et al 2013). Metagenomic sequencing, although comprehensive, can be expensive, requires extensive computational resources, and may be limited by the lack of reference databases for accurate annotation of resistance genes (Gupta et al 2020). High-throughput sequencing, in particular, has significantly advanced our ability to capture the complexity of microbial communities and resistance elements in WWTP influent, providing a more holistic view of the AMR burden (Xu et al 2023).

In recent years, prioritization of AMR risks in environmental matrices has become possible with the development of ranking systems (Martinez et al 2015). For example, the Zhang framework (Zhang et al., 2021) allows for a systematic evaluation of the potential risk posed by various ARGs, considering factors such as their prevalence, mobility, and association with pathogens. This ranking approach is particularly useful in guiding environmental monitoring efforts and mitigating the spread of resistance by identifying the most concerning ARGs present in WWTP influent (Garner et al 2024). Other frameworks such as Goh et al (2022), and Pruden et al (2018) also provide valuable approaches for assessing AMR risks in environmental matrices. Although the risk-based AMR assessment frameworks offer valuable information concerning AMR in environmental reservoirs, they might require extensive datasets to quantify exposure and health impacts, which can be challenging to obtain in diverse environmental settings. For example, the databases might be incomplete or biased towards well-studied ARGs.

In this study, we propose a new index tested on WWTP influent, that integrates Zhang et al (2021)’s ranking framework. We demonstrate how this new Resistance Persistence Index (RPI) works and propose it as a valuable tool for future studies on AMR in environmental settings.

## 2. Materials and Methods

Wastewater influent samples (500 ml) were collected from a local WWTP during both wet and dry seasons, once a week for four weeks. Collected samples were transported on ice to the Anand Lab at Hensill Hall, San Francisco State University and stored at -20°C until DNA extraction. Wastewater influent samples were processed following the microbe-phage wastewater DNA/RNA concentration and extraction protocol outlined in protocols.io (Rasile et al 2024). This protocol involved the concentration of viral and microbial DNA/RNA using a series of filtration and extraction steps designed to optimize recovery of both phage and microbial genetic material. The DNA extracts were then sent to the University of California, San Diego (UCSD) for 16S rRNA amplicon sequencing and shotgun sequencing, which produced an average of approximately 15 million reads per sample. The 16S rRNA data were analyzed using the Tourmaline bioinformatics pipeline (as per Thompson et al 2022). The shotgun sequencing results were analyzed using the NMDC Edge (Eloe-Fadrosh et al 2022) and Chan Zuckerberg ID (Kalantar et al 2020) platform to characterize the ARGs and microbial communities present in the samples.

ARG diversity and richness was calculated using the Shannon Diversity Index and Bray Curtis similarity index to compare across the wet and dry seasons. To determine the risk, persistence and mobility of ARGs, we introduce the Resistance Persistence Index (RPI) score by integrating the risk assessment outcomes from the Zhang framework (as per Zhang et al 2021) with persistence as per ARG abundance stability across seasons in wastewater influent. This approach allows a comprehensive assessment of AMR potential within WWTP influent.

Firstly, to apply the Zhang et al (2021) framework, we analyzed each ARG based on human association, gene mobility and pathogenic association. We checked if ARGs are abundant in wet and dry season samples, as human-associated environments like wastewater may show enrichment in one season and not both, and determined if ARGs are linked to high-risk pathogens, such as ESKAPE pathogens (e.g., *Klebsiella, Staphylococcus, Acinetobacter*). We assessed ARG mobility by identifying mobile elements (e.g., plasmids, integrons) associated with each ARG. In this manner, risk categories (1, 2, 3, 4) were classified based on a combination of ARG prevalence, association with pathogens, and potential for human exposure as per Zhang et al (2021). Rank 1 (current threats) were human-associated, mobile ARGs found in pathogens, and considered the highest risk. Rank 2 (future threats) were human-associated and mobile but not yet pathogens. Rank 3 were human-associated but not mobile, and rank 4 was low-risk ARGs, neither human-associated nor mobile.

Secondly, Persistence scores were determined using the following formula:

Persistence Score = 1 - |RPM_dry - RPM_wet| / (RPM_dry + RPM_wet + ε)

where RPM represents the relative persistence measure for dry and wet seasons, and ε is a small constant (e.g., 10^^-9^) to avoid division by zero. This formula evaluates the stability of ARGs by comparing their persistence across different environmental conditions, with higher scores indicating ARGs that are stable and resistant to environmental changes.

Thirdly, Mobility score was calculated as the proportion of mobile ARGs within each family: Mobility Score=Number of Mobile ARGs in Family/Total Number of ARGs in Family

This score ranges from 0 to 1, where 1 indicates all ARGs in the family are mobile

To analyze ARG mobility persistence by season, we identified mobile ARGs in each season based on indicators (e.g., plasmid, integron), summarized the number and abundance of mobile ARGs for both wet and dry seasons and compared the patterns in ARG mobility to see if there are differences in the abundance of mobile elements between seasons. This can help compare mobile ARG driven seasonal shifts in composition (e.g., higher diversity or abundance of mobile ARGs during the wet season over the dry season), which might help in predicting periods of higher resistance risk. Mobility scores are critical as they can indicate a high potential for horizontal gene transfer.

Therefore, the Resistance Persistence Index (RPI) was calculated by integrating the risk, persistence, and mobility scores. The RPI score provides a comprehensive assessment of each ARG’s risk level, taking into account its potential to persist and spread in the environment.

Risk-Persistence Index RPI=(Risk Rank Score)+(Persistence Score)+(Mobility Score)

Risk Rank Score = Convert each of Zhang et al (2021)’s ranks to a numerical scale (Rank I = 4, Rank II = 3, Rank III = 2, Rank IV = 1).

Persistence Score = average persistence score for each ARG or family

Mobility score = Each ARG gets assigned a score of 0 or 1 with 1 being mobile and 0 being not mobile. Following this the average mobility score is calculated for every ARG family.

All statistics are based on contigs of size >= 500 bp, unless otherwise noted (e.g., “# contigs (>= 0 bp)” and “Total length (>= 0 bp)” include all contigs).Contig statistics were generated for the metagenomic assemblies to provide an overview of the sequencing data quality and assembly completeness. A total of 1,040,144 contigs were assembled, with 284,834 contigs being 1,000 bp or longer and 9,176 contigs exceeding 5,000 bp. The largest contig was 228,270 bp. The total length of contigs ≥ 500 bp was approximately 1.05 billion base pairs. The N50 value, representing the contig length at which 50% of the total assembly length is contained, was 1,027 bp, while the N90 value was 563 bp. The GC content of the assembled sequences was calculated to be 52.36%. Data on raw reads is available at NMDC Edge.

## Results

A general analysis of the 16S rRNA dataset revealed alpha and beta diversity across all samples and provided a broad picture of diversity, highlighting both within-sample diversity (alpha; Figure 1-left) and between-sample differences (beta; Figure 1-right, bottom). The richness values and Shannon index for both wet and dry seasons appear high and similar. The PCoA plot displays two distinct clusters for wet and dry seasons demonstrating certain species being more prevalent in one season over the other. A closer examination of the influent samples demonstrated that pathogen prevalence varied notably by season. Pathogens, particularly focus pathogens, e.g., *Klebsiella, Pseudomonas*, etc. were found to be significantly more prevalent in the wet season compared to the dry season.

**Figure 1.**
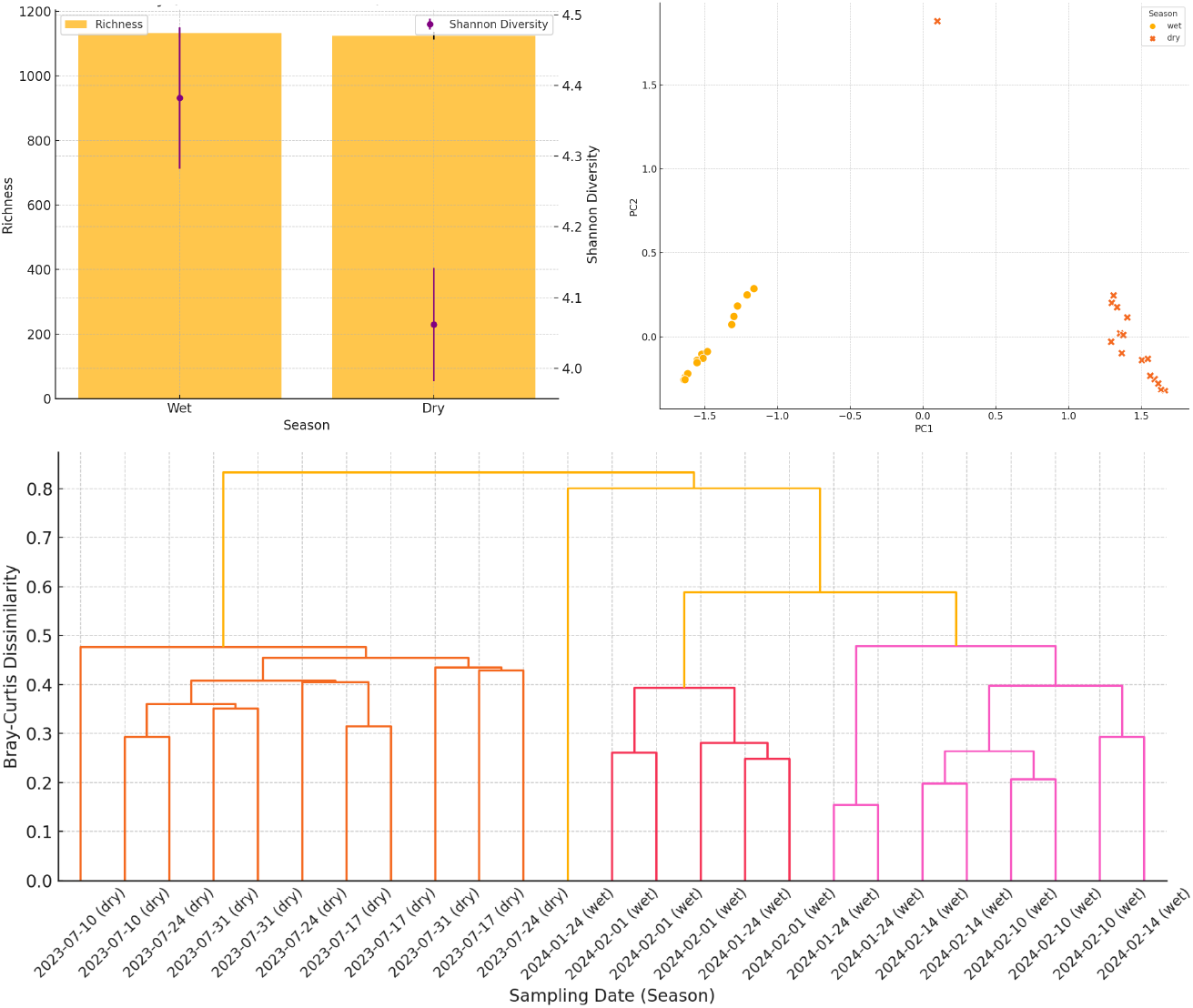
Left - 16S rRNA Seasonal diversity (richness and shannon index) with confidence intervals. Right - PCoA plot of 16S rRNA beta diversity (Bray-Curtis) by season; Bottom - Hierarchical clustering of samples based on beta diversity, the dendrogram shows the relationships between the samples

Understanding the dynamics of resistance in relation to environmental and clinical factors is critical for developing effective interventions to combat AMR. The insights gathered from these analyses can inform public health strategies, guide antibiotic stewardship programs, and shape future research on antimicrobial resistance.

We found 707 unique resistance genes in the dry season and 1064 unique resistance genes in the wet season. When we analyzed the distribution of resistance mechanisms across the samples and seasons (Table 1), “Antibiotic Inactivation” was the most prevalent resistance mechanism in both seasons, but significantly more so in the wet season. Calculating the average read counts per gene and their variability by season resulted in 190 gene counts (6.68 +/- 9.95) in the dry season when compared with 560 gene counts (16.31 +/- 88.53) in the wet season. When we conducted a correlation analysis between total reads and reads per million (RPM) for the resistance genes, the correlation coefficient (-0.0051, p<0.05) indicated no significant linear relationship. With reference to the ARG diversity from the shotgun sequencing dataset, there was distinct hierarchical clustering of AMR genes, as evidenced by a heatmap of the 25 most abundant AMR genes in the influent (Figure 3).

**Table 1.** Summary of resistance mechanism counts.

| Resistance Mechanism | Wet | Dry |
| --- | --- | --- |
| Antibiotic inactivation | 1830 | 687 |
| Antibiotic efflux | 971 | 383 |
| Target alteration | 387 | 198 |
| Target protection | 306 | 142 |
| Target replacement | 105 | 40 |

**Figure 3.**
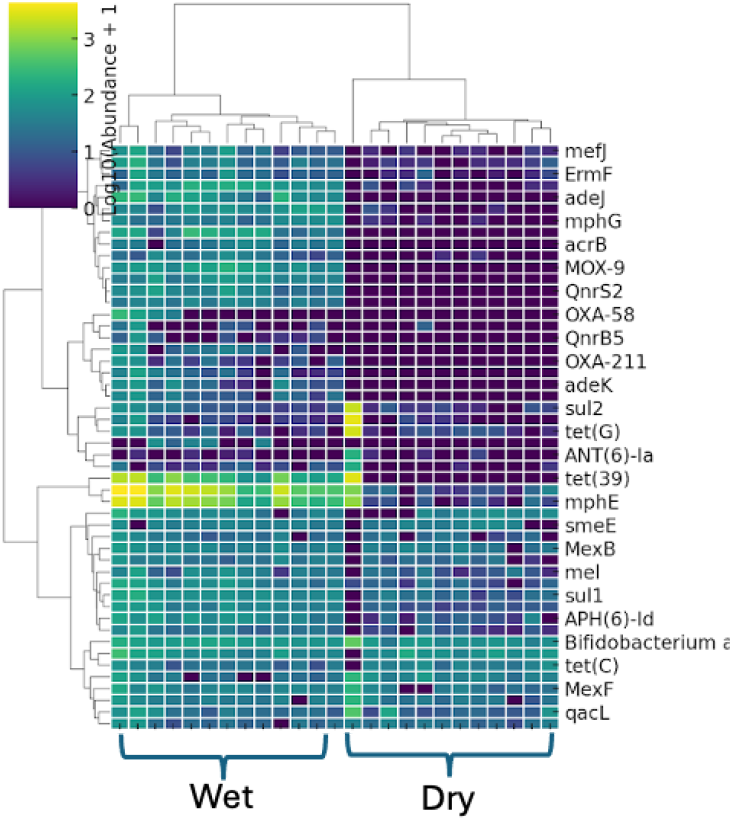
Hierarchical clustering of the 25 most abundant AMR genes in wastewater influent

To further contextualize these findings, we introduce a new Risk-Persistence Index (RPI) framework that integrates risk assessment with microbial persistence and mobility. This framework allows for a nuanced understanding of how ARGs interact within the ecosystem and provides a tool for predicting the implications of microbial resistance in varying environmental conditions.

First we ranked the ARG as per Zhang et al (2021) to risk level (Rank I to IV) based on human association, mobility, and pathogenic association (Figure 4). Next, we calculated a persistence (seasonal stability) and mobility score for each ARG family and calculated an RPI score (scatter plot in Figure 5). This allows us to visually compare where ARG families would conventionally sit versus their position in the integrated Risk-Persistence Index (RPI). The Rank I ARGs stand out because they are few in number (12 unique ARGs) but have a relatively high abundance (Average RPM 7.67) when compared to Rank III (412 unique ARGs; average RPM 6.42), and Rank IV (1249 unique ARGs; average RPM 0.23) with the exception of Rank II (102 unique ARGs; average RPM 30.3). This indicates that, although limited in diversity, these ARGs are highly prevalent within the samples. Such a pattern is typical of high-risk ARGs that are well adapted to thrive in human-impacted environments, where selective pressures, like antibiotic use, may favor their persistence. Next, the visualization of RPI versus other metrics helps to assess the combined effect of persistence and mobility on the risk associated with different ARG families, revealing how the RPI is influenced by these factors.

**Figure 4.**
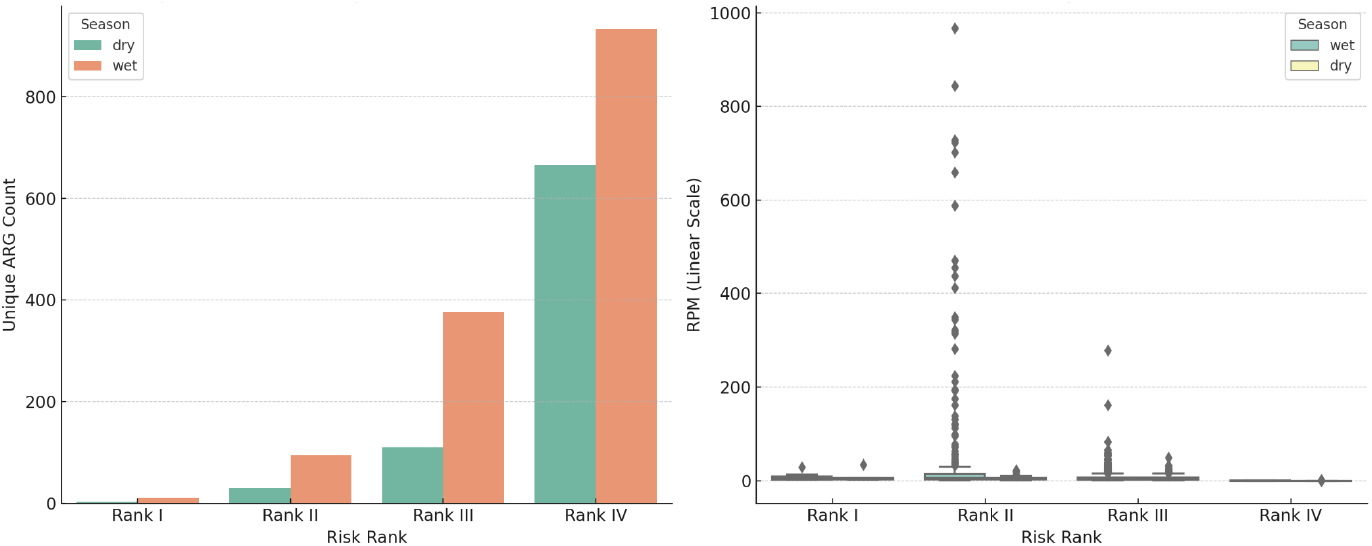
Unique ARG counts (left) and ARG abundance, rpm (right) by season and by risk rank

**Figure 5.**
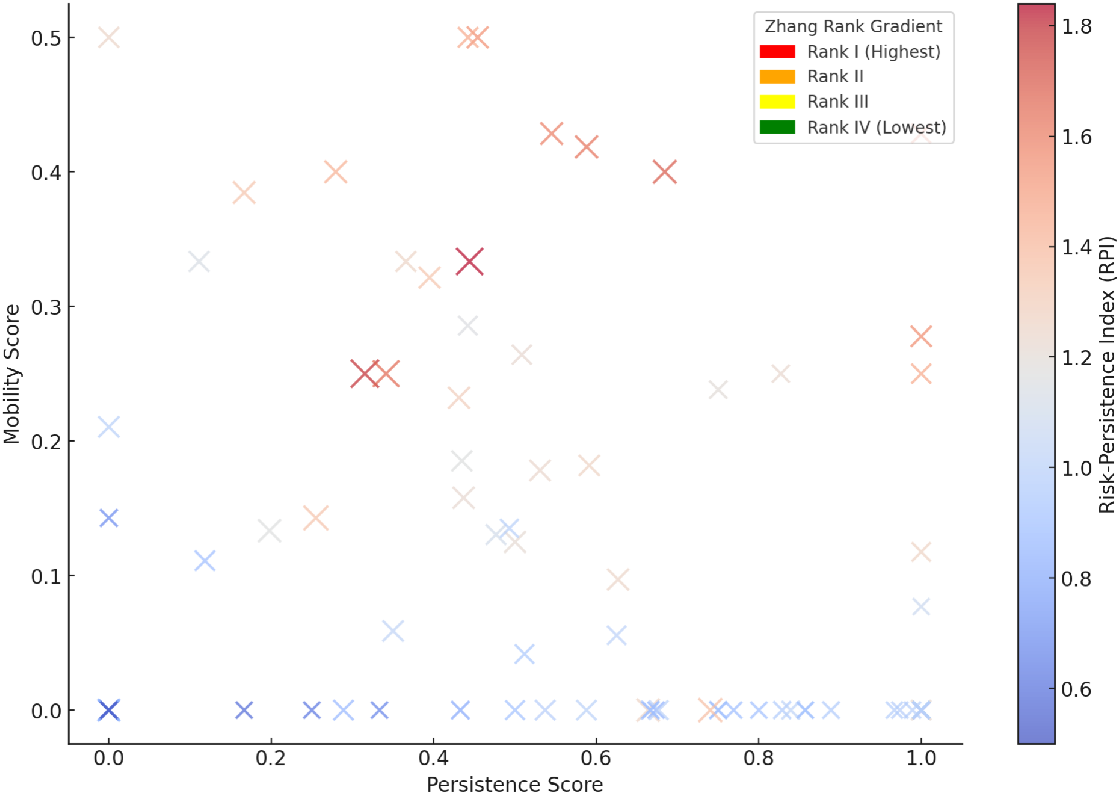
ARG family by persistence (seasonal stability), mobility, risk rank (as per Zhang et al 2021) and integrated RPI score (this study). Point size reflects Risk Rank (larger for higher risk), RPI color gradient indicates combined risk impact.

The distribution of ranks suggests that families with higher mobility and persistence scores may not always be the highest risk, emphasizing the complexity of ARG dynamics.

## Discussion

Overall, from the 16S rRNA community analysis and shotgun dataset, the alpha diversity metrics indicate a stable microbial community across seasons in terms of species richness and evenness. The beta diversity analysis highlights a significant seasonal shift in community composition, with specific taxa and ARGs likely driving the observed differences. These results suggest that environmental factors significantly influence microbial community structures despite overall stability in diversity metrics. Moreover, the higher prevelance of pathogens in the wet over the dry season suggests that seasonal variations may influence the dynamics of pathogen populations in the influent, possibly due to increased runoff associated with atmospheric river events.

The notable increase in the number of unique resistance genes during the wet season (1064) compared to the dry season (707) suggests that environmental factors in the wet season may favor the proliferation of a wider variety of resistance genes. Further investigation into the conditions during each season may help identify specific factors contributing to this difference. Additionally, the wet season showed higher counts across all resistance mechanisms compared to the dry season, suggesting that the conditions during the wet season may be conducive to greater resistance expression or gene transfer among microbial communities. Lastly, the average number of reads per gene was significantly higher in the wet season compared to the dry season. This suggests that there is a greater abundance of these resistance genes during this period. The standard deviation in the wet season also indicated a wider variability in read counts, with some genes having an exceptionally high number of reads (up to 1629), which could signify specific resistance genes that are highly prevalent under wet conditions.

We hypothesize that the higher average and variability during the wet season may indicate a robust microbial community that is potentially more resilient to antibiotic pressure. The lack of significant linear relationship between the total reads and RPM suggests that the abundance of specific resistance genes (as indicated by RPM) does not necessarily correlate with the total microbial load in the samples. It may imply that resistance genes can exist independently of the total biomass or that their expression can vary regardless of the total reads.

The risk ranking framework revealed a relatively lower number of unique ARGs (e.g., 12) in Rank 1 - which show the highest risk yet may be less diverse. The high average RPM suggests these ARGs are abundantly expressed where they occur, highlighting a concentrated but high-impact threat. The presence of Rank I ARGs in both seasons, especially in “wet” (human-impacted) samples, reinforces their potential public health impact, suggesting areas for targeted monitoring and intervention. Rank II recorded more unique ARGs (e.g., 102), with a higher average RPM compared to other ranks, indicating ARGs that may spread easily due to their mobility. The presence of mobile but currently non-pathogenic ARGs hints at a reservoir that could transfer into pathogenic hosts over time, especially if ecological conditions favor ARG dissemination. With a large number of unique ARGs (e.g., 412), Rank III shows a diverse profile, though with relatively low average abundance (RPM).

Rank III ARGs are prevalent but less threatening due to their lack of mobility and pathogenic linkage. Their diversity suggests they may be under selective pressure in human-associated environments, potentially becoming more mobile or pathogenic through horizontal gene transfer over time. Lastly, Rank IV contains the highest diversity of ARGs (e.g., 1249 unique types), but with very low average RPM, indicating that these ARGs are widespread but inactive or less influenced by anthropogenic factors. These ARGs might represent a background resistance gene pool in the environment. While they pose little immediate threat, Rank IV ARGs could evolve under selective pressures in wastewater or other human-impacted sites.

The integrated Risk-Persistence Index (RPI) approach revealed that some Rank II ARG families could indeed exhibit higher overall risk than Rank I families when factors like persistence and mobility are considered. Although the risk ranking primarily focuses on pathogenicity and mobility, by adding seasonal persistence and nuanced mobility scores, we captured a fuller risk picture. Some Rank II families, for instance, might have high persistence and mobility, which could elevate their practical risk over certain Rank I families with lower seasonal stability or mobility. This finding suggests that traditional risk ranks might miss critical aspects of ARG behavior in environmental contexts, emphasizing the importance of the RPI for comprehensive risk assessment.

## Conclusion

The introduction of the Resistance Persistence Index (RPI) enhances our ability to assess the risk of antimicrobial resistance in wastewater environments by integrating persistence, mobility, and risk ranking. Our findings highlight significant seasonal variations in ARG prevalence, with a notable increase in diversity and abundance during wet conditions. By employing a comprehensive approach that considers both ecological dynamics and human impact, the RPI serves as a valuable framework for predicting resistance risks and informing public health strategies. The results underscore the need for targeted monitoring and management of high-risk ARGs in WWTPs, particularly during wet seasons, to mitigate the spread of antimicrobial resistance in environmental settings.

## Data Availability

All sequence data are deposited on NMDC Edge and available upon request as well.

## Acknowledgments

We are grateful for the research funding from the DOE BRaVE Phage Foundry, graduate student support from San Francisco State University’s Student Enrichment Opportunities Office and postbac scholar support from NSF Bay Area RAMP program in microbiome science. We are grateful to the WWTP officials and operations manager for sample retrieval.

